# A Machine-Learning Approach to Finding Gene Target Treatment Options for Long COVID

**DOI:** 10.1101/2025.02.07.25321856

**Authors:** Alejandro Lopez-Rincon

## Abstract

Long COVID, also known as post-acute sequelae of SARS-CoV-2 infection (PASC), encompasses a range of symptoms persisting for weeks or months after the acute phase of COVID-19. These symptoms, affecting multiple organ systems, significantly impact the quality of life. This study employs a machine-learning approach to identify gene targets for treating Long COVID. Using datasets GSE275334, GSE270045, and GSE157103, Recursive Ensemble Feature Selection (REFS) was applied to identify key genes associated with Long COVID. The study highlights the therapeutic potential of targeting genes such as PPP2CB, SOCS3, ARG1, IL6R, and ECHS1. Clinical trials and pharmacological interventions, including dual antiplatelet therapy and anticoagulants, are explored for their efficacy in managing COVID-19-related complications. The findings suggest that machine learning can effectively identify biomarkers and potential therapeutic targets, offering a promising avenue for personalized treatment strategies in Long COVID patients.

## 1. Introduction

Long COVID, also known as post-acute sequelae of SARS-CoV-2 infection (PASC), refers to a range of symptoms that persist for weeks or months after the acute phase of a COVID-19 infection has resolved. These symptoms can affect multiple organ systems, including the respiratory, cardiovascular, neurological, and gastrointestinal systems, significantly impacting the quality of life of those affected. Individuals with Long COVID may experience ongoing issues such as fatigue, shortness of breath, chest pain, joint pain, and cognitive difficulties like brain fog. The condition is associated with immune system dysregulation, chronic inflammation, and potential autoimmune reactions. Recognizing and diagnosing Long COVID is crucial to providing appropriate care and support for those affected, as the prolonged symptoms can severely affect daily activities and overall well-being [1].

It is estimated that 31%-69% of COVID-19 survivors will experience long COVID symptoms after initial recovery from SARS-CoV-2 infection [2]. Fatigue, cognitive issues (commonly known as brain fog), and post-exertional malaise (PEM) are frequently mentioned symptoms. However, there are over 200 symptoms that have been linked to Long COVID. The most prevalent symptom of long COVID worldwide is fatigue [3]. Additionally, cognitive-related symptoms may develop at later stages. Long COVID symptoms have been documented in both hospitalized and non-hospitalized individuals, with approximately 30% of non-hospitalized patients reporting lingering symptoms two months after initial infection [2].

In the realm of pharmacotherapy, several drugs are being explored for the treatment of long COVID. These include Anhydrous enol-oxaloacetate [4], Atorvastatin [5], Ivermectin [6], Methylnicotinamide [7], Montelukast [8], and N115 sodium pyruvate nasal spray [9]. Additionally, combinations such as Naltrexone with nicotinamide adenine dinucleotide [10], and nasal sprays composed of local corticosteroids and antihistamines [11], are under investigation. Another notable treatment is S-1226 [12], which is a mixture of aerosolized carbon dioxide and nebulized perflubron.

Given the diverse range of symptoms associated with long COVID, most treatment trials have focused on non-pharmacological approaches. These trials aim to address multiple symptoms simultaneously or specifically target respiratory issues. Interventions often include therapies designed to reduce inflammation and enhance tissue oxygenation. Additionally, some trials explore methods to improve overall cardiovascular health and support the body’s natural healing processes. By targeting these underlying mechanisms, researchers hope to alleviate the persistent and varied symptoms experienced by long COVID patients [13].

Considering the global incidence of long COVID, there is a significant amount of omics data available. Using machine learning (ML), we aim to identify biomarkers that could serve as gene targets. ML has been previously utilized for gene-targeted therapies, such as in [14], where machine learning models using drug-induced gene expression profiles were employed to predict targets for diseases like hypertension, diabetes mellitus, rheumatoid arthritis, and schizophrenia. By applying these models to gene perturbation data, researchers prioritized targets based on their therapeutic potential. Similarly, in [15], the authors proposed a novel machine learning model that integrates genomic data to predict tumor drug sensitivity more accurately. Using statistical methods and Random Forests, the model preprocesses gene expression data, selects significant genomic features, and classifies drug sensitivity. The findings demonstrate that this approach significantly improves prediction accuracy for both targeted and non-specific chemotherapy drugs, potentially expanding the application of molecular targeted therapies and enhancing personalized cancer treatment.

In [16], the article discusses the use of machine learning (ML) to identify biomarkers that can predict patient responses to treatments. These biomarkers are crucial for personalized medicine, as they help tailor treatments to individual patients based on their unique biological characteristics. The section highlights how ML models can analyze complex, high-dimensional data from various sources, such as genomics, proteomics, and clinical data, to identify patterns and correlations that traditional methods might miss. By leveraging these advanced techniques, researchers can improve the accuracy of predictions, leading to better treatment outcomes and more efficient drug development processes.

## 2. Methods

In previous studies, we used Recursive Ensemble Feature Selection (REFS) for *omics* studies. REFS is an ensemble method for feature selection, programmed in python designed to enhance reproducibility in the biomarker discovery field across different datasets to compensate for batch error [17, 18, 19]. It consists of eight classifiers (Stochastic Gradient Descent on linear models, SVM, Gradient Boosting, Random Forest, Logistic Regression, Passive Aggressive classifier, Ridge Classifier and Bagging) from the scikit-learn toolbox [20] that rank the features at each iteration. REFS employs a nested approach within a 10-fold cross-validation scheme to produce robust and unbiased results [21]. REFS has been used successfully for feature selection in *omics* data, e.g., miRNA [22, 23], mRNA [24, 25, 26], and microbiome [27, 28, 29, 17, 18, 19]. Compared to other methods, REFS has proven to be more effective in feature selection than using Univariate Feature Selection (UFS) [30], Recursive Feature Elimination (RFE) [31], Elastic Net (EN) [32], Genetic Algorithms (GALGO) [33], Least Absolute Shrinkage and Selection Operator (LASSO) [34] and Ensemble Feature Selection with Complete Linear Aggregation (EFS-CLA) [35].

Unlike other methodologies such as GRACES and DNP, which do not automatically determine the optimal number of features (*k*), REFS is capable of finding a suitable *k* automatically. This capability is why we selected REFS to first identify and select the number of features. To validate our analysis we use 10-fold cross-validation [21] and we report with a classifier that was not part of the ensemble to avoid *over-fitting* [36], Multi-layer Perceptron (MLP) classifier. Then, using the results of the discovery dataset we apply the found genes to the other 2 datasets. In this case, our discovery dataset is GSE275334, and the test datasets are GSE270045 and GSE157103 (Fig. 1). Next, using *Open Targets* [37] and *DrugBank* [38], we identify known drugs, diseases and pharmacogenetics to discover treatment options. Finally, we look for clinical trials, that validate our targets [39].

**Figure 1:**
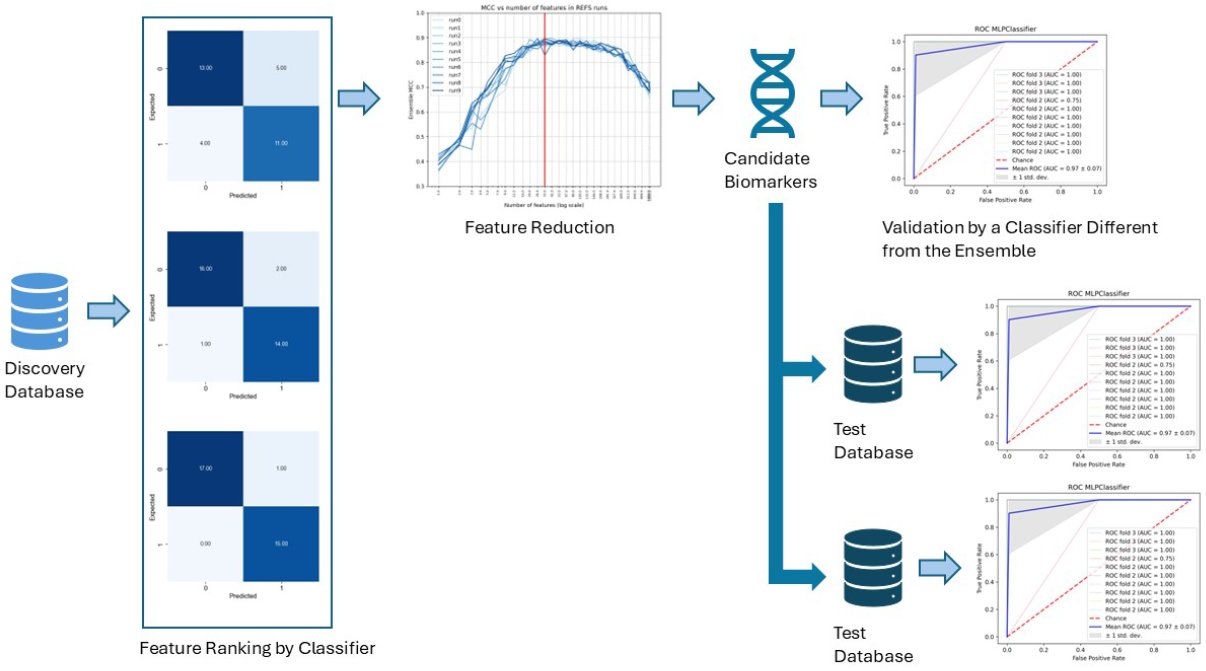
Methodology to Validate our Biomarkers.

### 2.1. GSE275334

The dataset GSE275334 [40], “Immune Exhaustion in ME/CFS and long COVID” involves gene expression analysis using the NanoString nCounter Immune Exhaustion gene expression panel. This panel includes 785 genes to investigate mechanisms behind T cell, B cell, and NK cell exhaustion in disease. RNA was extracted from peripheral blood mononuclear cells (PBMCs) from participants with ME/CFS (n=14), long COVID (n=15), and healthy controls (n=18). The study aimed to compare gene expression profiles between these groups to understand immune exhaustion in ME/CFS and long COVID. The data was normalized against controls to account for background noise and platform variation, and differential expression was reported between ME/CFS, long COVID, and healthy controls. As discovery set, we only used long COVID patients (n=15) and healthy controls (n=18).

### 2.2. GSE270045

The dataset GSE270045 [41], “Upregulation of olfactory receptors and neuronal-associated genes highlights complex immune and neuronal dysregulation in Long COVID patients” involves gene expression analysis using high throughput sequencing. RNA was isolated from the whole blood of 19 patients who developed myalgic encephalomyelitis/chronic fatigue syndrome (ME/CFS) following acute SARS-CoV-2 infection, and from 17 healthy controls (HCs). The study aimed to investigate the differential gene expression patterns between these groups to understand the immune and neuronal dysregulation in long COVID patients.

### 2.3. GSE157103

The dataset GSE157103 [42], “Largescale Multi-omic Analysis of COVID-19 Severity” involves a comprehensive systems analysis using RNA-seq and high-resolution mass spectrometry. The study analyzed 128 plasma and leukocyte samples from hospitalized patients with or without COVID-19 (n=102 and 26, respectively) and with varying degrees of disease severity. The aim was to generate abundance measurements for over 17,000 transcripts, proteins, metabolites, and lipids, and compile them with clinical data into a curated relational database. As validation set, we compared ICU (n=50) against non-ICU COVID patients (n=50).

## 3. Results

Using the raw counts from dataset GSE275334 and samples for long COVID (n=15), and healthy controls (n=18), we applied REFS and reduced it from 635 to 7 genes (Fig. 2).

**Figure 2:**
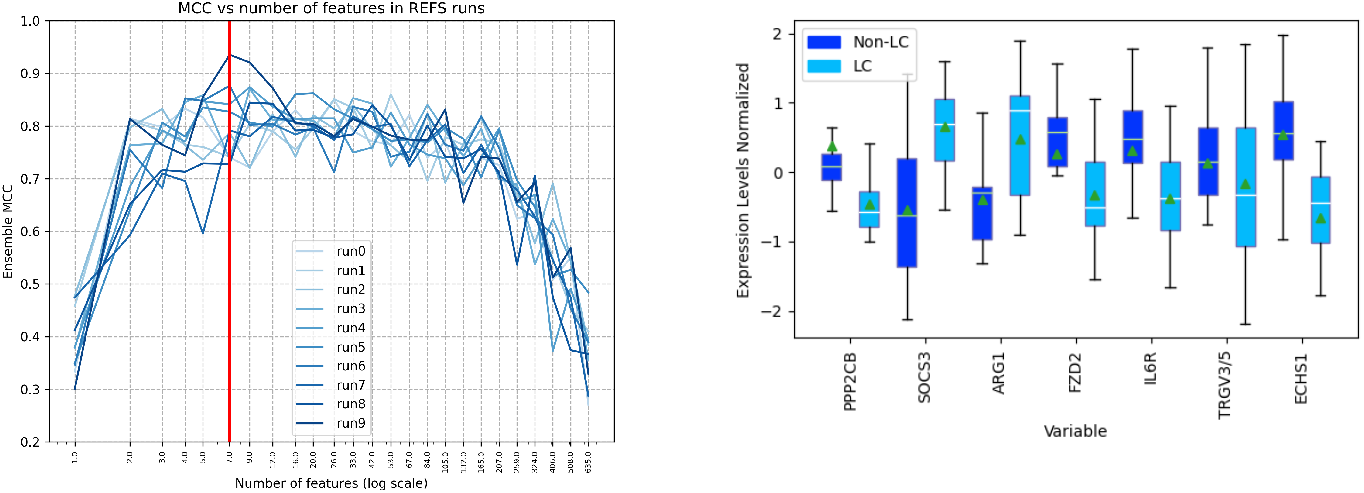
(Left) Reduction of the genes from 635 to 7. (Right) Box-plot comparison of the 7 genes.

Then, we calculate the ROC curve using the MLP classifier, a classifier not part of the ensemble, to avoid over-fitting, and compared it to *SelecKBest* using F-score metric (Fig. 3).

**Figure 3:**
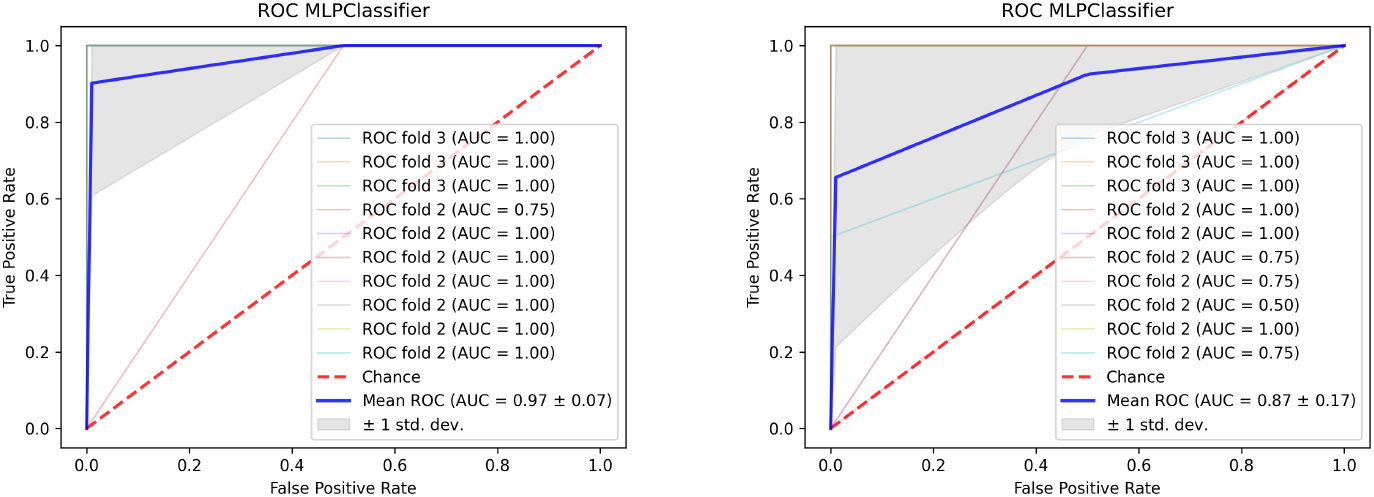
(Left) AUC using MLP 10-fold cross-validation of the 7 genes found with REFS. (Right) AUC using F-Score as a metric, *k* = 7.

Then, we validate the found genes in 2 extra datasets using MLP classifier; GSE270045 (Fig. 4) and GSE157103(Fig. 5).

**Figure 4:**
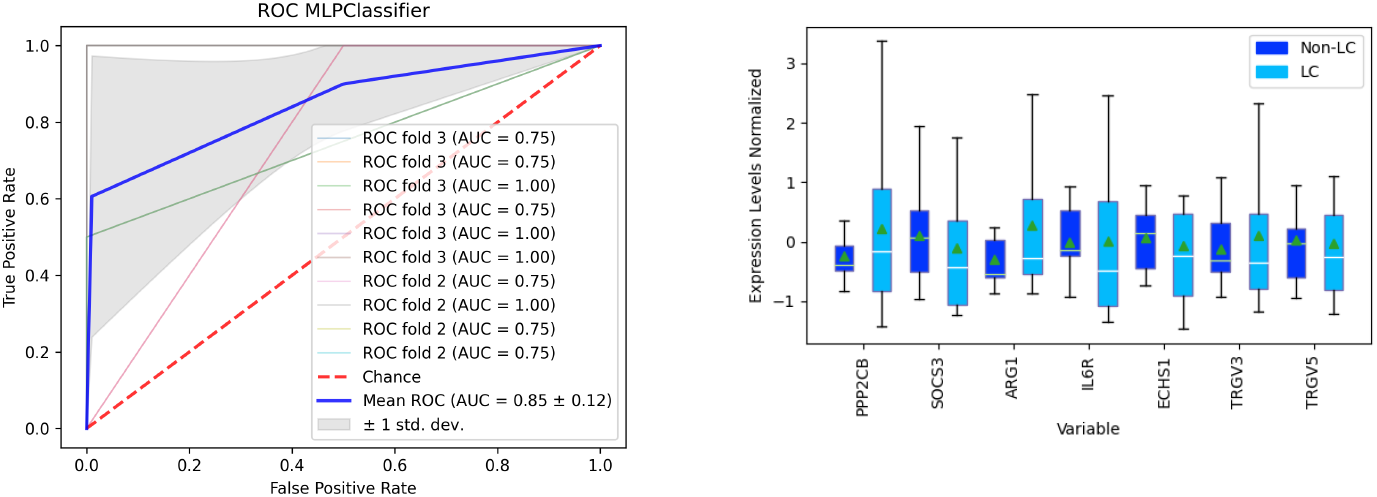
(Left) AUC using MLP 10-fold cross-validation of dataset GSE270045 with the available genes, 6 of 7. (Right) Box-plot of the genes.

**Figure 5:**
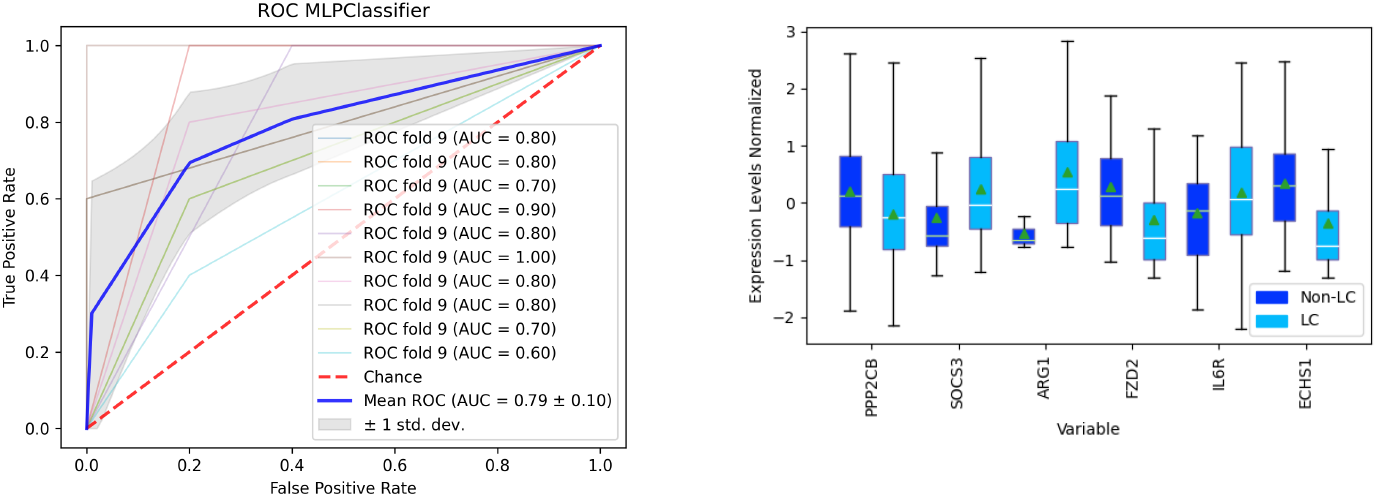
(Left) AUC using MLP 10-fold cross-validation of dataset GSE270045 with the available genes, 6 of 7. (Right) Box-plot of the genes.

## 4. Discussion and Conclusion

Following the AUC mean from Figs. 4 and 5, we get a diagnostic accuracy of very good and good respectively [43] in the test datasets using the found genes from the discovery dataset. From the 7 found genes, 5 of them are connected to COVID. From these 5, 4 of them are on clinical trials (Table 1):

**Table 1:** Clinical trials, with drugs that target the found genes.

| Target Gene | Known Drug | Clinical Trials in COVID-19/Long COVID (Max 5) |
| --- | --- | --- |
| PPP2CB |  |  |
| SOCS3 | Resveratrol | NCT05601180, NCT04400890, NCT04799743, NCT04542993, NCT04666753 |
| ARG1 | Bitolterol, albuterol | NCT04681079 |
| FZD2 |  |  |
| IL6R | Tocilizumab | NCT04317092, NCT04363736, NCT04924829, NCT04479358, NCT04331795, |
| TRGV3/5 |  |  |
| ECHS1 | Clopidogrel+Aspirin | NCT04333407, NCT04368377, NCT04518735, |

Connected to COVID-19 in recent studies.
Gene Target drugs in clinical trials for COVID-19

- **PPP2CB**: Recent studies have highlighted the therapeutic potential of pulmonary mesenchymal stem cells (MSCs) in treating severe COVID-19 and its pulmonary sequelae. MSCs from severe cases show upregulation of genes related to cell dispersion, migration, and the interferon-stimulated response (ISR), while downregulating NF-*κ*B upstream receptors, suggesting a role in mitigating the cytokine storm [44]. Many NF-*κ*B inhibitors, such as PPP2CB, OPTN, NFKBIA, and FHL2, were notably upregulated, indicating that MSCs are not involved in the cytokine storm seen in severe COVID-19 cases. This suggests that lung MSCs detect immune threats and respond protectively within the pulmonary environment, reinforcing their potential as a therapeutic option in cell-based treatments for COVID-19. Additionally, the PPP2CB gene, encoding a subunit of protein phosphatase 2A, is implicated in modulating immune responses and inflammation, making it a potential target for therapeutic interventions in severe COVID-19 cases [45].
- **SOCS3**: Flavonoids, such as flavone and naringenin, have shown promising therapeutic potential in various health conditions. Flavone has been found to induce the expression of the suppressor of cytokine signaling 3 (SOCS3) gene, which plays a role in neurodegenerative diseases [46]. Additionally, flavonoids are suggested as critical nutritional supplements for alleviating and shortening the period associated with post-COVID-19 syndrome [47]. Recent studies have highlighted the palliative effects of naringenin and naringin on various COVID-19 sequelae [48]. Furthermore, vitamin D and resveratrol have emerged as powerful immunomodulatory agents and enzymatic inhibitors in the battle against SARS-CoV-2 infection [49].
- **ARG1**: Selective Beta 2-adrenergic Agonists, such as bitolterol, are primarily used to dilate air passages in the lungs that have become narrowed due to disease or inflammation. This makes them effective in treating conditions like asthma and chronic obstructive pulmonary disease (COPD). Recent studies have explored the potential of drug repurposing, utilizing molecular docking procedures to identify bitolterol as a promising inhibitor of COVID-19 protease [50]. Emerging evidence from preliminary clinical results suggests that bronchodilators, including bitolterol, may offer significant benefits when repurposed for treating pneumonia or bronchospasm associated with COVID-19. This dual functionality of bronchodilators, providing both anti-inflammatory and bronchodilating effects, positions them as a potential therapeutic strategy for managing COVID-19 symptoms [51].
- **IL6R**: The IL6R gene, encoding the interleukin-6 receptor, is crucial in regulating immune response and inflammation. Clinical trials such as NCT04317092 [52], NCT04363736 [53], NCT04924829 [54], NCT04479358 [55], and NCT04331795 [56] explore IL-6 receptor inhibitors for conditions like severe COVID-19, rheumatoid arthritis, and colorectal cancer, aiming to reduce inflammation and improve patient outcomes. Additionally, these studies investigate genetic variants to predict treatment efficacy and toxicity. Emerging evidence also highlights IL-6’s role in long COVID, where elevated levels contribute to prolonged inflammation. Targeting the IL6R gene may help alleviate long-term symptoms, underscoring the therapeutic potential of IL-6 receptor inhibitors in both acute and chronic COVID-19 management [57, 58].
- **ECHS1** : ECHS1 (Enoyl-CoA Hydratase, Short Chain 1) deficiency, a mitochondrial disorder, can complicate the management of COVID-19 due to its impact on metabolic processes. Patients with ECHS1 deficiency and neurodegenerative disease may benefit from a treatment regimen that includes dual antiplatelet therapy with Clopidogrel and Aspirin. Clinical trials such as NCT04333407 [59], NCT04368377 [60], and NCT04518735 [61] have explored the efficacy of such therapies in managing COVID-19-related complications. These studies highlight the potential to improve outcomes by preventing cardiac issues, reducing hypoxemia, and assessing the benefits of various anticoagulant regimens. Additionally, research has shown that targeting these pathologies with anticoagulant therapy can help resolve symptoms and improve patient outcomes. This approach aims to mitigate the severe effects of COVID-19 in patients with underlying metabolic disorders, enhancing their overall prognosis [62].

In conclusion, this study demonstrates the efficacy of using machine learning, specifically Recursive Ensemble Feature Selection (REFS), to identify key gene targets associated with Long COVID. The identified genes, including PPP2CB, SOCS3, ARG1, IL6R, and ECHS1, exhibit significant therapeutic potential. These genes are involved in critical biological processes such as immune response modulation and inflammation regulation, making them promising targets for therapeutic interventions. The study also highlights ongoing clinical trials and pharmacological interventions, such as dual antiplatelet therapy and anticoagulants, which are being explored for their effectiveness in managing Long COVID symptoms. These interventions aim to reduce inflammation, improve cardiovascular health, and support the body’s natural healing processes. The findings underscore the potential of machine learning in advancing personalized treatment strategies for Long COVID patients, paving the way for more targeted and effective therapies. Overall, this research contributes to a deeper understanding of Long COVID and offers a promising avenue for developing personalized and effective therapeutic approaches to improve the quality of life for those affected by this condition.

## Data Availability Statement

All data produced are available online at: https://github.com/steppenwolf0/longCOVIDTarget

## References

[1] A. Raveendran, R. Jayadevan, S. Sashidharan, Long covid: an overview, Diabetes & Metabolic Syndrome: Clinical Research & Reviews 15 (3) (2021) 869–875.

[2] H. C. Koc, J. Xiao, W. Liu, Y. Li, G. Chen, Long covid and its management, International journal of biological sciences 18 (12) (2022) 4768.

[3] Centers for Disease Control and Prevention, Long covid or post-covid conditions, accessed: 2025-02-03 (2024). URL long-covid-signs-symptoms

[4] A. Cash, D. L. Kaufman, Oxaloacetate treatment for mental and physical fatigue in myalgic encephalomyelitis/chronic fatigue syndrome (me/cfs) and long-covid fatigue patients: a non-randomized controlled clinical trial, Journal of translational medicine 20 (1) (2022) 295.

[5] I.-S. Investigators, et al., Atorvastatin versus placebo in patients with covid-19 in intensive care: randomized controlled trial, bmj 376 (2022).

[6] C. Fernández-de Las-Peñas, J. Torres-Macho, J. A. Catahay, R. Macasaet, J. V. Velasco, S. Macapagal, M. Caldararo, B. M. Henry, G. Lippi, A. Franco-Moreno, et al., Is antiviral treatment at the acute phase of covid-19 effective for decreasing the risk of long-covid? a systematic review, Infection 52 (1) (2024) 43–58.

[7] M. Chudzik, M. Burzyńska, J. Kapusta, Use of 1-mna to improve exercise tolerance and fatigue in patients after covid-19, Nutrients 14 (15) (2022) 3004.

[8] F. Mera-Cordero, S. Bonet-Monne, J. Almeda-Ortega, A. García-Sangenís, O. Cunillera-Puértolas, S. Contreras-Martos, G. Alvarez-Muñoz, R. Monfà, M. Balanzo-Joué, R. Morros, et al., Double-blind placebo-controlled randomized clinical trial to assess the efficacy of montelukast in mild to moderate respiratory symptoms of patients with long covid: E-speranza covid project study protocol, Trials 23 (1) (2022) 19.

[9] C. R. Lupfer, R. Nadler, R. Amen, A. Martin, Inhalation of sodium pyruvate to reduce the symptoms and severity of respiratory diseases including covid-19, long covid, and pulmonary fibrosis, Eur J Respir Med 3 (3) (2021) 229–237.

[10] A. Isman, A. Nyquist, B. Strecker, G. Harinath, V. Lee, X. Zhang, S. Zalzala, Low-dose naltrexone and nad+ for the treatment of patients with persistent fatigue symptoms after covid-19, Brain, Behavior, & Immunity-Health 36 (2024) 100733.

[11] T. K. Dietz, K. N. Brondstater, Long covid management: a mini review of current recommendations and underutilized modalities, Frontiers in Medicine 11 (2024) 1430444.

[12] A. A. Antar, A. L. Cox, Translating insights into therapies for long covid, Science translational medicine 16 (773) (2024) eado2106.

[13] F. Ceban, A. Leber, M. Y. Jawad, M. Yu, L. M. Lui, M. Subramaniapillai, J. D. Di Vincenzo, H. Gill, N. B. Rodrigues, B. Cao, et al., Registered clinical trials investigating treatment of long covid: a scoping review and recommendations for research, Infectious Diseases 54 (7) (2022) 467–477.

[14] K. Zhao, Y. Shi, H.-C. So, Prediction of drug targets for specific diseases leveraging gene perturbation data: a machine learning approach, Pharmaceutics 14 (2) (2022) 234.

[15] R. Miao, H.-H. Chen, Q. Dang, L.-Y. Xia, Z.-Y. Yang, M.-F. He, Z.-F. Hao, Y. Liang, Beyond the limitation of targeted therapy: improve the application of targeted drugs combining genomic data with machine learning, Pharmacological Research 159 (2020) 104932.

[16] J. Vamathevan, D. Clark, P. Czodrowski, I. Dunham, E. Ferran, G. Lee, B. Li, A. Madabhushi, P. Shah, M. Spitzer, et al., Applications of machine learning in drug discovery and development, Nature reviews Drug discovery 18 (6) (2019) 463–477.

[17] D. Rojas-Velazquez, S. Kidwai, A. D. Kraneveld, A. Tonda, D. Oberski, J. Garssen, A. Lopez-Rincon, Methodology for biomarker discovery with reproducibility in microbiome data using machine learning, BMC bioinformatics 25 (1) (2024) 26.

[18] L. N. Peralta-Marzal, D. Rojas-Velazquez, D. Rigters, N. Prince, J. Garssen, A. D. Kraneveld, P. Perez-Pardo, A. Lopez-Rincon, A robust microbiome signature for autism spectrum disorder across different studies using machine learning, Scientific Reports 14 (1) (2024) 814.

[19] D. Rojas-Velazquez, S. Kidwai, T. C. Liu, M. A. El-Yacoubi, J. Garssen, A. Tonda, A. Lopez-Rincon, Understanding parkinson’s: The microbiome and machine learning approach, Maturitas 193 (2025) 108185.

[20] F. Pedregosa, G. Varoquaux, A. Gramfort, V. Michel, B. Thirion, O. Grisel, M. Blondel, P. Prettenhofer, R. Weiss, V. Dubourg, et al., Scikit-learn: Machine learning in python, the Journal of machine Learning research 12 (2011) 2825–2830.

[21] A. Vabalas, E. Gowen, E. Poliakoff, A. J. Casson, Machine learning algorithm validation with a limited sample size, PloS one 14 (11) (2019) e0224365.

[22] A. Lopez-Rincon, M. Martinez-Archundia, G. U. Martinez-Ruiz, A. Schoenhuth, A. Tonda, Automatic discovery of 100-mirna signature for cancer classification using ensemble feature selection, BMC bioinformatics 20 (2019) 1–17.

[23] A. Lopez-Rincon, L. Mendoza-Maldonado, M. Martinez-Archundia, A. Schönhuth, A. D. Kraneveld, J. Garssen, A. Tonda, Machine learningbased ensemble recursive feature selection of circulating mirnas for cancer tumor classification, Cancers 12 (7) (2020) 1785.

[24] S. Kidwai, P. Barbiero, I. Meijerman, A. Tonda, P. Perez-Pardo, P. Lio, A. H. van der Maitland-Zee, D. L. Oberski, A. D. Kraneveld, A. Lopez-Rincon, A robust mrna signature obtained via recursive ensemble feature selection predicts the responsiveness of omalizumab in moderate-to-severe asthma, Clinical and Translational Allergy 13 (11) (2023) e12306.

[25] P. I. Metselaar, L. Mendoza-Maldonado, A. Y. F. Li Yim, I. Abarkan, P. Henneman, A. A. Te Velde, A. Schönhuth, J. A. Bosch, A. D. Kraneveld, A. Lopez-Rincon, Recursive ensemble feature selection provides a robust mrna expression signature for myalgic encephalomyelitis/chronic fatigue syndrome, Scientific reports 11 (1) (2021) 4541.

[26] D. Rojas-Velazquez, A. Tonda, I. Rodriguez-Guerra, A. D. Kraneveld, A. Lopez-Rincon, Multi-objective evolutionary discretization of gene expression profiles: Application to covid-19 severity prediction, in: J. Correia, S. Smith, R. Qaddoura (Eds.), Applications of Evolutionary Computation, Springer Nature Switzerland, Cham, 2023, pp. 703–717.

[27] K. Kamphorst, A. Lopez-Rincon, A. M. Vlieger, J. Garssen, E. van’t Riet, R. M. van Elburg, Predictive factors for allergy at 4–6 years of age based on machine learning: A pilot study, PharmaNutrition 23 (2023) 100326.

[28] M. Benner, A. Lopez-Rincon, S. Thijssen, J. Garssen, G. Ferwerda, I. Joosten, R. G. van der Molen, A. Hogenkamp, Antibiotic intervention affects maternal immunity during gestation in mice, Frontiers in Immunology 12 (2021) 685742.

[29] J. Blankestijn, A. Lopez-Rincon, A. Neerincx, S. Vijverberg, S. Hashimoto, M. Gorenjak, O. Sardón-Prado, P. Corcuera, J. Korta-Murua, M. Pino-Yanes, et al., Classifying asthma control using salivary and fecal microbiome in children with moderate to severe asthma: results from the syspharmpedia study (2022).

[30] A. V. Lazo, P. Rathie, On the entropy of continuous probability distributions (corresp.), IEEE Transactions on Information Theory 24 (1) (1978) 120–122.

[31] I. Guyon, J. Weston, S. Barnhill, V. Vapnik, Gene selection for cancer classification using support vector machines, Machine learning 46 (2002) 389–422.

[32] A. Sokolov, D. E. Carlin, E. O. Paull, R. Baertsch, J. M. Stuart, Pathway-based genomics prediction using generalized elastic net, PLoS computational biology 12 (3) (2016) e1004790.

[33] V. Trevino, F. Falciani, Galgo: an r package for multivariate variable selection using genetic algorithms, Bioinformatics 22 (9) (2006) 1154– 1156.

[34] R. Tibshirani, Regression shrinkage and selection via the lasso, Journal of the Royal Statistical Society Series B: Statistical Methodology 58 (1) (1996) 267–288.

[35] T. Abeel, T. Helleputte, Y. Van de Peer, P. Dupont, Y. Saeys, Robust biomarker identification for cancer diagnosis with ensemble feature selection methods, Bioinformatics 26 (3) (2010) 392–398.

[36] X. Ying, An overview of overfitting and its solutions, in: Journal of physics: Conference series, Vol. 1168, IOP Publishing, 2019, p. 022022.

[37] G. Koscielny, P. An, D. Carvalho-Silva, J. A. Cham, L. Fumis, R. Gasparyan, S. Hasan, N. Karamanis, M. Maguire, E. Papa, et al., Open targets: a platform for therapeutic target identification and validation, Nucleic acids research 45 (D1) (2017) D985–D994.

[38] C. Knox, M. Wilson, C. M. Klinger, M. Franklin, E. Oler, A. Wilson, A. Pon, J. Cox, N. E. Chin, S. A. Strawbridge, et al., Drugbank 6.0: the drugbank knowledgebase for 2024, Nucleic acids research 52 (D1) (2024) D1265–D1275.

[39] D. A. Zarin, T. Tse, R. J. Williams, R. M. Califf, N. C. Ide, The clinicaltrials. gov results database—update and key issues, New England Journal of Medicine 364 (9) (2011) 852–860.

[40] N. Eaton-Fitch, P. Rudd, T. Er, L. Hool, L. Herrero, S. Marshall-Gradisnik, Immune exhaustion in me/cfs and long covid, JCI insight 9 (20) (2024) e183810.

[41] A. Rahmati, S. Shahbaz, M. Osman, J. W. C. Tervaert, S. Elahi, Blood transcriptomic analyses do not support sars-cov-2 persistence in patients with post-covid-19 condition with chronic fatigue syndrome, The Lancet Microbe (2024).

[42] K. A. Overmyer, E. Shishkova, I. J. Miller, J. Balnis, M. N. Bernstein, T. M. Peters-Clarke, J. G. Meyer, Q. Quan, L. K. Muehlbauer, E. A. Trujillo, et al., Large-scale multi-omic analysis of covid-19 severity, Cell systems 12 (1) (2021) 23–40. [43]

[43] A.-M. Šimundić, Measures of diagnostic accuracy: basic definitions, ejifcc 19 (4) (2009) 203.

[44] A. Henriques-Pons, D. G. Beghini, V. d. S. Silva, S. Iwao Horita, F. A. B. d. Silva, Pulmonary mesenchymal stem cells in mild cases of covid-19 are dedicated to proliferation; in severe cases, they control inflammation, make cell dispersion, and tissue regeneration, Frontiers in Immunology 12 (2022) 780900.

[45] L. Yao, J. Li, Y. Li, P. Wang, J. Ma, Q. Tu, Y. Yuan, Y. Chen, L. Wang, Y. Chen, et al., Regγ regulates antiviral response by activating tbk1-ifnβ signaling through degradation of ppp2cb (2024).

[46] D. Ochoa, A. Hercules, M. Carmona, D. Suveges, J. Baker, C. Malangone, I. Lopez, A. Miranda, C. Cruz-Castillo, L. Fumis, et al., The next-generation open targets platform: reimagined, redesigned, rebuilt, Nucleic acids research 51 (D1) (2023) D1353–D1359.

[47] A. Bardelčíková, A. Miroššay, J. Šoltýs, J. Mojžiš, Therapeutic and prophylactic effect of flavonoids in post-covid-19 therapy, Phytotherapy Research 36 (5) (2022) 2042–2060.

[48] S. Liu, M. Zhong, H. Wu, W. Su, Y. Wang, P. Li, Potential beneficial effects of naringin and naringenin on long covid—a review of the literature, Microorganisms 12 (2) (2024) 332.

[49] C. Russo, M. S. Valle, L. Malaguarnera, I. R. Romano, L. Malaguarnera, Comparison of vitamin d and resveratrol performances in covid-19, Nutrients 15 (11) (2023) 2639.

[50] S. Aishwarya, K. Gunasekaran, R. S. Jansi, G. Sangeetha, From genomes to molecular dynamics–a bottom up approach in extrication of sars cov-2 main protease inhibitors, Computational Toxicology 18 (2021) 100156.

[51] Y. Yu, B. Fang, X.-D. Yang, Y. Zheng, One stone two birds: antiinflammatory bronchodilators as a potential pharmacological strategy for covid-19, Frontiers in Pharmacology 14 (2023) 1185076.

[52] F. Perrone, M. C. Piccirillo, P. A. Ascierto, et al., Tocilizumab in covid-19 pneumonia (tocivid-19), medRxiv (2020). URL https://clinicaltrials.gov/ct2/show/NCT04317092

[53] H.-L.R. Ltd, A phase ii, open-label, randomized, multicenter study to investigate the pharmacodynamics, pharmacokinetics, safety, and efficacy of 8 mg/kg or 4 mg/kg intravenous tocilizumab in patients with moderate to severe covid-19 pneumonia (2020). URL https://clinicaltrials.gov/ct2/show/NCT04363736

[54] M. Anaclara, Safety and effectiveness observational study of anti il-6 tocilizumab in hospital admitted patients with severe covid-19 pneumonia, Blood (2024). URL https://clinicaltrials.gov/ct2/show/NCT04924829

[55] V. Authors, Low-dose tocilizumab versus standard of care in hospitalized patients with covid-19 (covidose-2) (2023). URL https://clinicaltrials.gov/ct2/show/NCT04479358

[56] V. Authors, Tocilizumab to prevent clinical decompensation in hospitalized, non-critically ill patients with covid-19 pneumonitis (covidose) (2020). URL https://clinicaltrials.gov/ct2/show/NCT04331795

[57] J.-X. Yin, Y. L. Agbana, Z.-S. Sun, S.-W. Fei, H.-Q. Zhao, X.-N. Zhou, J.-H. Chen, K. Kassegne, Increased interleukin-6 is associated with long covid-19: a systematic review and meta-analysis, Infectious Diseases of Poverty 12 (1) (2023) 43.

[58] X. Wang, G. Tang, Y. Liu, L. Zhang, B. Chen, Y. Han, Z. Fu, L. Wang, G. Hu, Q. Ma, et al., The role of il-6 in coronavirus, especially in covid-19, Frontiers in Pharmacology 13 (2022) 1033674.

[59] ClinicalTrials.gov, Preventing cardiac complication of covid-19 disease with early acute coronary syndrome therapy: A randomised controlled trial, ClinicalTrials.gov (2020). URL https://clinicaltrials.gov/ct2/show/NCT04333407

[60] ClinicalTrials.gov, Enhanced platelet inhibition treatment improves hypoxemia in patients with severe covid-19 and hypercoagulability: A case control, proof of concept study, ClinicalTrials.gov (2020). URL https://clinicaltrials.gov/ct2/show/NCT04368377

[61] ClinicalTrials.gov, Evolution of covid-19 in anticoagulated or antiaggregated patients (corona study), ClinicalTrials.gov (2020). URL https://clinicaltrials.gov/ct2/show/NCT04518735

[62] G. Laubscher, A. Khan, C. Venter, J. Pretorius, D. Kell, E. Pretorius, Treatment of long covid symptoms with triple anticoagulant therapy, Research Square (2023). doi:10.21203/rs.3.rs-2697680/v1. URL https://europepmc.org/article/PPR/PPR633963

